# An MRI-based Deep Learning Model to Predict Parkinson’s Disease Stages

**DOI:** 10.1101/2021.02.19.21252081

**Authors:** Azadeh Mozhdehfarahbakhsh, Saman Chitsazian, Prasun Chakrabarti, Tulika Chakrabarti, Babak Kateb, Mohammad Nami

## Abstract

Parkinson’s disease (PD) is amongst the relatively prevalent neurodegenerative disorders with its course of progression classified as prodromal, stage1, 2, 3 and sever conditions. With all the shortcomings in clinical setting, it is often challenging to identify the stage of PD severity and predict its progression course. Therefore, there appear to be an ever-growing need need to use supervised and unsupervised artificial intelligence and machine learning methods on clinical and paraclinical datasets to accurately diagnose PD, identify its stage and predict its course. In today’s neuro-medicine practices, MRI-related data are regarded beneficial in detecting various pathologies in the brain. In addition, the field has recently witnessed a growing application of deep learning methods in image processing often with outstanding results. Here, we applied Convolutional Neural Networks (CNN) to propose a model helping to distinguish different stages of PD. The results showed that our current MRI-based CNN model may potentially be employed as a suitable method for the distinction of PD stages at a high accuracy rate (0.94).

## 1. Introduction

Parkinson’s Disease (PD) is a neurodegenerative disease more commonly observed in the elderly. Almost 1% of the world population suffer from PD of whom many cases present with complicated motor and cognitive issues. Over the course of the disease progression, cognitive and behavioral symptoms including a wide variety of personality changes, depressive disorders, memory dysfunction and emotion dysregulation may emerge. Moreover, movement symptoms get worse as the disease progresses. Dementia should be diagnosed at an early stage so that appropriate therapeutic interventions can be used to prevent cognitive deterioration.^1–3^

In routine practice, clinicians diagnose PD based on the presenting symptoms including slowness, stiffness, tremor, and balance/coordination difficulties. Yet, such symptoms and their progression rate may differ case by case. Currently, there seems to be no specific blood test or biomarker to accurately diagnose PD or monitor underlying changes as the condition escalates.^1^

Over the last three decades, MRI (magnetic resonance imaging) has been used as a tool to diagnose and differentiate various neurological diseases from suspected PD.^4^

Researchers have found that changes in disease progression can be detected by brain MRI under a special protocol. Such neuroimaging modalities could be utilized in clinical trials, as an objective way to monitor the effectiveness of treatments.^5^

With reference to the use of Convolutional Neural Networks (CNNs) in image processing, there appear to be new horizons to provide a comprehensive approach towards a wide variety of applications such as image recognition, segmentation, and retrieval. This new paradigm has created exceptional results over the recent years in terms of analyzing the content of images, speech and videos. Many studies have shown that the presenting state-of-the-art CNNs retains accuracies that surpass human-level performance.^6^

Moreover, feature representation has been one of the most important factors in medical image processing. Deep learning methods such as CNN extracts and uses new and hidden features which would not be considered by traditional machine learning methods.^7,8^

## 2. Background and Related Works

The present investigation has been an attempt to adopt CNN upon brain MRI of PD patients aiming to develop trained predictive models which could efficiently classify PD stages compared to the same dataset from healthy individuals.

Even though the severity of PD and its various stages play an important role in timely interventions, few studies proposed a model to predict and diagnose the severity of PD. However, there are numerous works to predict PD using various machine learning techniques. Srishti et al.^6^ proposed a deep neural network architecture for the prediction of PD severity on UCI’s Parkinson’s Telemonitoring Voice Dataset of patients. Franz *et al*.^9^ used a dataset of 8,661 minutes of IMU data from 30 patients, and defined the motor state (off, on, dyskinetic) based on MDS-UPDRS global bradykinesia item as well as the AIMS upper limb dyskinesia item. Having used a 1-minute window size as an input for a CNN trained model on the data from a subset of patients, they achieved a three-class balanced accuracy of 0.654 on data from previously unseen subjects. They have used serum samples from a clinically well-characterized longitudinally followed Michael J Fox Foundation cohort of PD patients to aid the prediction of PD progression using machine learning models.

Similarly, in a survey by Das et al.^11^, they examined different classification techniques in diagnosing the PD, among other machine learning techniques such as regression and decision tree. Their findings indicated neural network as a preferred classifier. In some other research works, features extracted from speech signals^12,13^ was used for predicting the severity of PD. Genain et al. (2014) used Bagged decision trees to predict the severity of PD from voice recordings of patients and found a 2% improvement in the prediction accuracy level. Moreover, Malek et al. (2015) used 40-features dataset and achieved 9 best features using Local Learning Based Feature Selection (LLBFS) to classify PD subjects according to their UPDRS score into four classes (Healthy, Early, Intermediate and Advance). Other than the above investigations, Cole et al.^14^ used the data collected from wearable sensors and shown that dynamic machine learning algorithms could be used to detect the severity of tremors and Dyskinesia. In addition, Angeles et al.^15^ innovated a wearable sensor system to record movement of their arms to identify changes of their performance during Deep Brain Simulation Therapy.

In line with the above works, Nilashi et al.^16^ suggested a new hybrid intelligent system using Adaptive neuro fuzzy inference system (ANFIS) and Support Vector Regression (SVR) for predicting the PD progression. Liu et al.^17^ provided a system by means of PCA for feature extraction and Fuzzy KNN for classification and PD diagnostic. Likewise, Polat et al.^18^ employed the Fuzzy C-Means (FCM) clustering method and KNN to propose a system to diagnose PD. Some other works were also done to design a PD prediction system using parallel feed forward Neural Network after which compared against a rule-based system to propose the decision model^19^. Li et al.^20^ suggested a fuzzy based nonlinear transformation method where PCA was used for feature extraction and SVM to predict the progressive course of PD. Another proposed model was a hybrid intelligent system using clustering, feature reduction and classification methods aiming to accurately diagnose PD^21^.

## 3. Research Methodology

### 3.1 DATASET acquisition

The PPMI dataset which was applied in this report (Parkinson’s Progression Markers Initiative, RRID:SCR_006431) is related to a clinical investigation to verify progression markers in PD (https://www.ppmi-info.org/). The acquired images from the PPMI database were obtained 4.5h after the injection of 111 to 185 MBq of DaTSCAN.

A total of 100 images from PPMI database were used in our analyses. Specifically, our study analyzed the baseline acquisition from 20 subjects for each stage of PD and 20 normal controls (NC).

### 3.2. Proposed Methodology

The proposed methodology towards predicting the severity of PD was based on deep learning. At first, the PD patients’ MRI data were collected and normalized using the min-max normalization. The data subsequently underwent pre-processing. In the next step, deep neural networks were designed with an input layer, hidden layers and an output layer.

### 3.3 CNNs for Classification

According to Figure 1 the scheme of CNN reveals 9 layers consisting five convolutional layers, the flatten layer, two fully connected layers and output layer. The output layer was in fact the network classifier layer which used the Soft Max function. Eventually, the number of outputs in this layer and the number of classes in the network training section were five. The convolutional layers used five 2D-kernels of [3×3] to sweep over the input topologies and transform them into feature maps. In addition, Stride of (2) and padding were employed with the convolution for the purpose of keeping the output feature maps with the size of the input. Additionally, the ReLu function was employed for the nonlinear activation function of the convolutional layers as well as fully connected layers. The output layer contained 5 neurons corresponding to the five disease condition. Classes comprised the Control, Prodromal, as well as stages 1, 2 and 3. The normalized data was then fed into the constructed deep neural network for the purpose of training and testing. The last layer yielded the prediction probability using the Soft Max activation (Figure 1).

**Figure 1.**
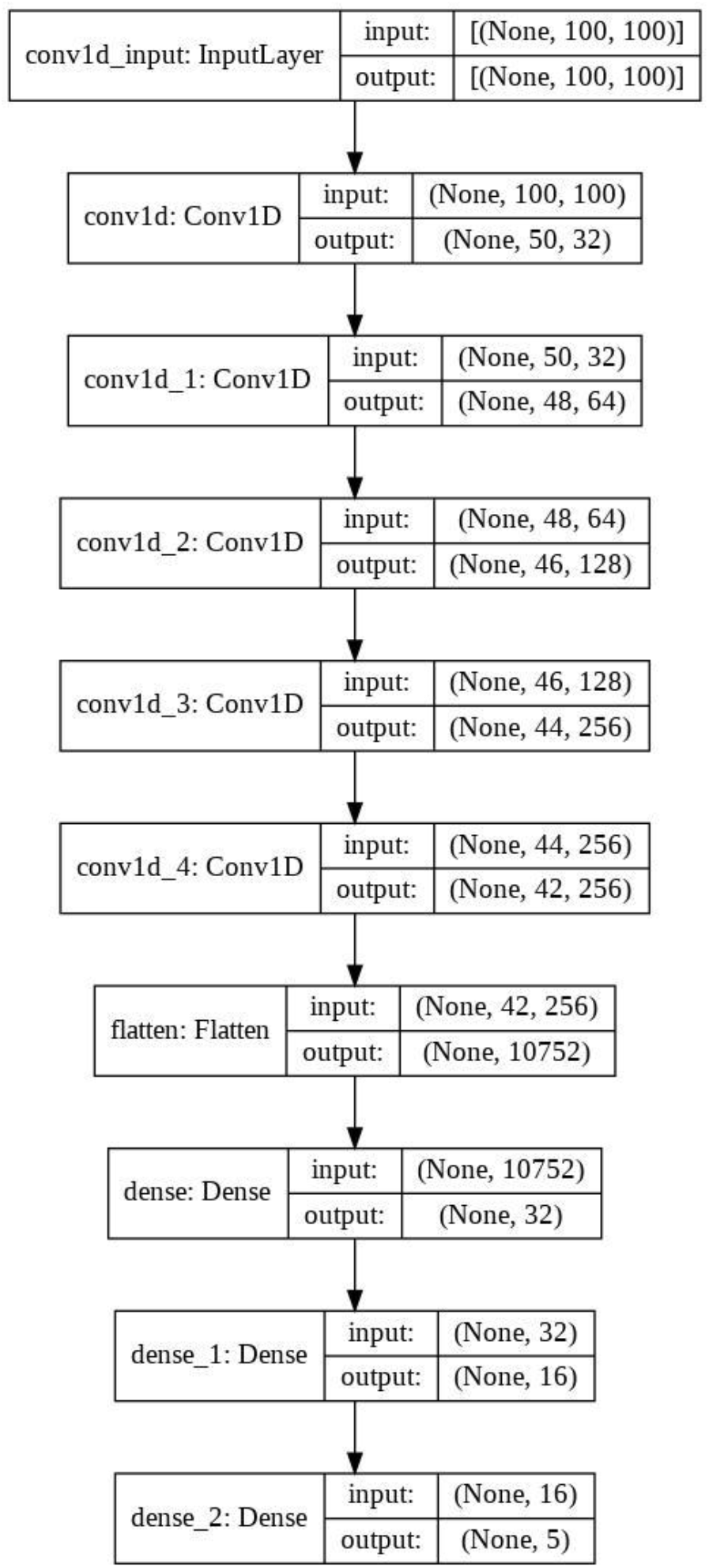
Summary of CNN model for classification of PD

### 3.4 K-fold Cross Validation

In order to test and generalize the deep learning results, we implemented cross validation to estimate how accurately our predictive model would perform in practice. The K-fold cross validation tool was simple to use, and complete data was conviniently used for the purpose “training and validation”. Our approach here applied the “K values of 10” in CNN method to diminish bias. Figure 3 gives the basics structure of “k-fold cross validation method” employed in our study (Figure 2).

**Figure 2.**
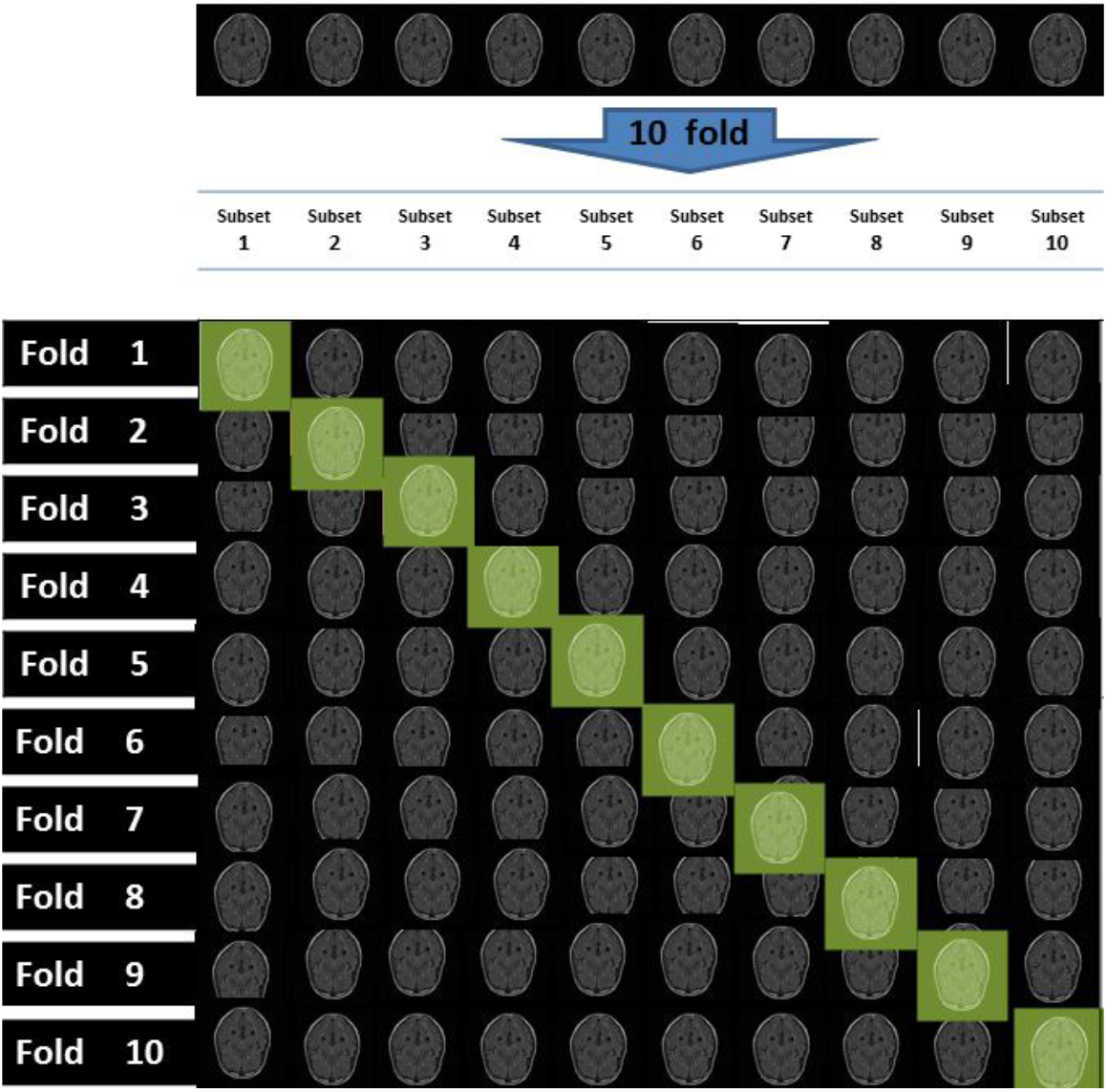
structure of 10-fold cross validation method

## 4. Results

The current proposed algorithm appeared to be potentially useful for the predicting the disease course in elderly PD patients. Artificial intelligence (AI) tools coupled with increasingly powerful biometric sensors would allow detecting abnormalities at prodromal and different stages of PD.

In the present work, our proposed algorithm was applied on PPMI dataset as described before. We also used Python programming language and the keras with tensorflow backend to implement the CNN.

### 4.1 Experimental Results of the method (CNN)

This section highlights the results of classification by CNN. Figures 3(a) and 3(b) indicate the progress of training in CNN model, and the corresponding fluctuations in accuracy and loss metrics. Different inquired evaluation criteria included the accuracy, loss, precision, recall and f1-score. 10-Fold CNN Mean for loss, accuracy after 60 epochs were 0.179, 0.94 respectively. Table 1 shows precision, recall and f1-score for all stages.

**Figures 4.**
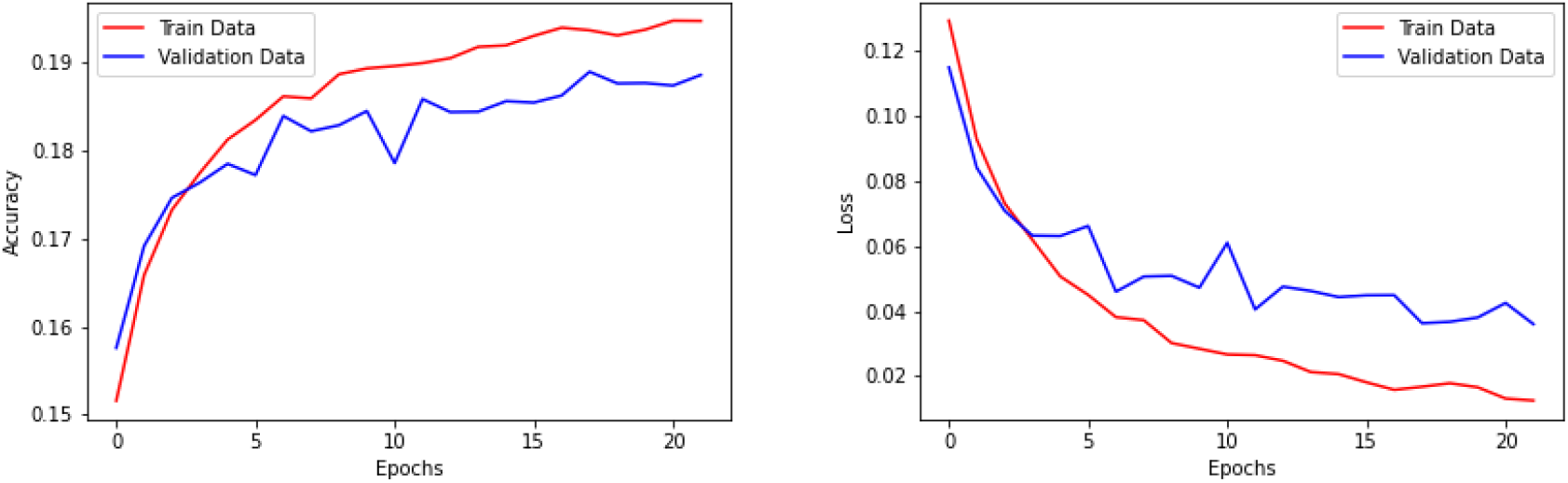
The averaged accuracy (a) and loss (b) of the model in 10-fold cross validation

**Table 1.** Metrics corresponding to all folds.

|  | precision | recall | f1-score | support |
| --- | --- | --- | --- | --- |
| 0 | 0.94 | 0.90 | 0.92 | 698 |
| 1 | 0.90 | 0.90 | 0.90 | 381 |
| 2 | 0.93 | 0.95 | 0.94 | 1457 |
| 3 | 0.88 | 0.86 | 0.87 | 484 |
| 4 | 0.98 | 0.98 | 0.98 | 1960 |

AUC/ROC or “Area under the ROC Curve” measures the classification model performance at various threshold settings. It indicates to what extend the model correctly stratifies the classes and distinguishes between them. Overall, higher AUC indicate that the model is genuinely found classes as they are. In our model, the higher the AUC, the better the model distinguishes different stages of PD and classes. This curve plots parameters including True Positive (TP), and False Positive (FP) Rates.

Figures 5a and 5b illustrate some of the main characteristics of the ROC curve. It simply plots TP vs. FP at different classes. Once the classification threshold diminished, our model could classify more samples as TP, whereas the increased threshold corresponded to a higher FP.

**Figure 5.**
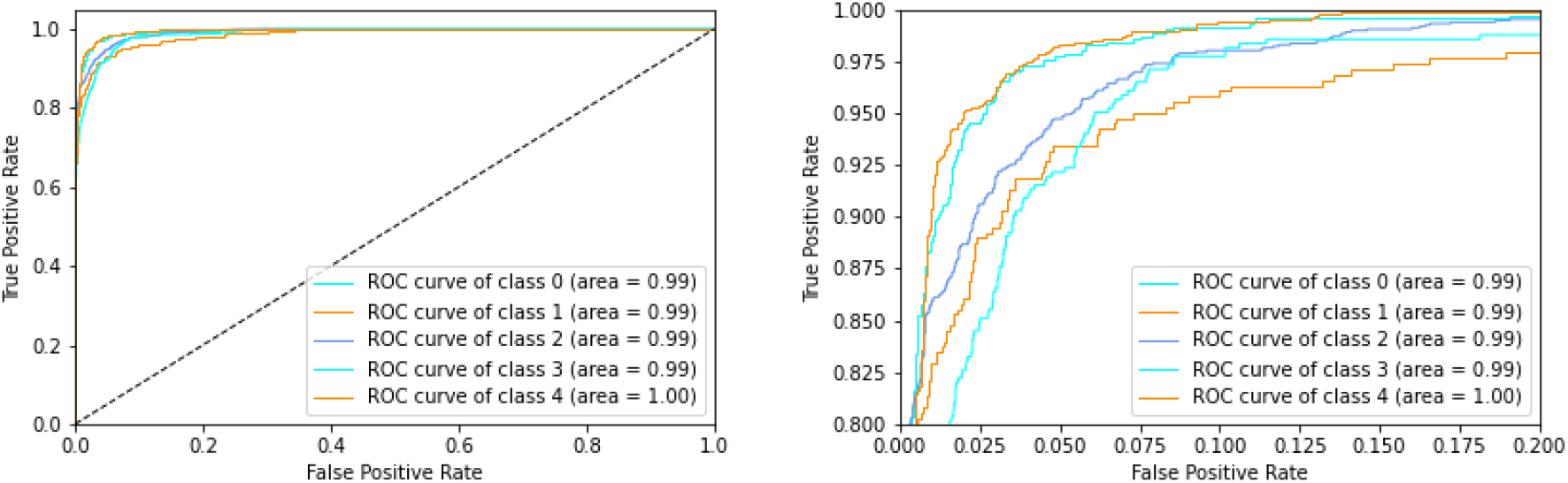
AUC or “Area under the ROC Curve.” measures the whole two-dimensional area underneath the ROC from (0,0) to (1,1).

## 5. Summary and Concluding Remarks

PD is one of the neurodegenerative disorders with its main symptoms detectable only after disease progression. Obviously, the interventions, medications and neuro-rehabilitation treatments should be appropriately selected as per the disease stage and on case-by-case basis. There are several problems with the mainstream clinical evaluation of PD at clinical setting. Those include human error and overseen clinical observations. To overcome such drawbacks in clinical evaluation, diagnosis and disease follow up in patients with AD, some studies have proposed methods towards the prediction of PD severity using machine learning methods on different datasets such as voice and UPDRS to distinguish the stages of the disease and healthy people.^6,9,10^ The present investigation used MRI data to diagnose stages of PD using deep neural networks.

Our results demonstrated that the proposed CNN model provides a high accuracy predictive accuracy (94%) with MRI data to distinguish healthy and PD as well as disease staging. Deep Neural Network classifiers are proposed for the detection of different stages of PD as compared to healthy controls datasets. This would expectedly improve the diagnosis and assist clinicians for timely intervention decision in patients with PD. Based on the present findings, CNN retains a proper capacity as a classifier over MRI data for different stages of PD at least in the context of the PPMI database.

## Data Availability

https://samsat.sums.ac.ir/page-Novin/fa/84/form/pId15008

